# Characterizing the Longitudinal Impact of Backward Locomotor Treadmill Training on Walking and Balance Outcomes in Chronic Stroke Survivors: A Randomized Single Center Clinical Trial

**DOI:** 10.1101/2024.09.11.24313519

**Authors:** Oluwole O. Awosika, Colin Drury, Amanda Garver, Pierce Boyne, Heidi J. Sucharew, Emily Wasik, Amit Bhattacharya, Kari Dunning, Pooja Khatri, Brett M. Kissela

**Author notes:** **Corresponding author**: Oluwole O. Awosika, MD, MS, 231 Albert Sabin Way, Medical Sciences Building, Room 7216, Cincinnati, OH 45267.

## Abstract

**Background and Purpose:** Walking and balance impairments after stroke are a global health concern, causing significant morbidity and mortality. However, effective strategies for achieving meaningful recovery in the chronic stages are limited. Backward locomotor treadmill training (BLTT) is a novel walking rehabilitation protocol that is safe, feasible, and likely beneficial in stroke survivors; however, its efficacy has not been tested. This single-center, randomized, assessor-blind clinical trial aims to test the preliminary efficacy of BLTT compared to forward locomotor treadmill training (FLTT) on walking speed, symmetry, and postural stability.

**Methods:** Forty stroke survivors [BLTT (N=19), FLTT (N=21); mean age= 56.3 ± 8.6 years; 53% Female; 30% Non-Hispanic Black] with mild-moderate walking impairment were enrolled. Participants underwent nine 30-minute BLTT or FLTT sessions over three weeks. The primary outcome was the mean change in the 10-meter walk test (10 MWT) at 24 hours post-training (24 hr POST). Secondary outcome measures were changes in spatiotemporal walking symmetry and postural stability during quiet standing at 24 hr POST. Retention was explored at Days 30- and 90 POST.

**Results:** We report clinically meaningful (≥ 0.16 m/s) improvements in overground walking speed at 24 hr POST, with retention up to Day 90 POST with BLTT and FLTT. However, contrary to our working hypothesis, no between-group differences in walking speed were observed. Nonetheless, we found that BLTT resulted in offline improvements in spatial symmetry and retention of subcomponents of the modified clinical test of sensory interaction on balance (mCTSIB), including the testing of proprio-vestibular integration up to Day 30 POST.

**Conclusion:** Among chronic stroke patients with mild-moderate walking impairment, BLTT and FLTT both resulted in long-lasting and clinically meaningful improvement in walking speed. However, preliminary findings suggest that BLTT may better comprehensively target walking asymmetry and sensory systems processing and integration.

## INTRODUCTION

Chronic walking and balance impairments after stroke are common and represent a significant cause of morbidity and mortality.^1^ While most stroke survivors can walk without continuous physical assistance from another person, relatively few have adequate walking speed to enable complete community independence. Additionally, stroke survivors are at an increased risk of falls compared to age-matched controls and often experience low balance confidence-- perpetuating a downward spiral of relative inactivity and secondary complications of stroke^2^ Consequently, the recovery of walking and balance are two of the most desired skills to regain with rehabilitation interventions.^3,4^

Walking and balance are physiologically distinct skills, yet they share many similarities from the motor control perspective.^5^ For example, both require multimodal processing to function efficiently, including cognitive, sensorimotor, postural, coordination, and musculoskeletal engagement.^3^ This multimodal processing may become impaired with aging and after neurologic injury such as stroke.^3^ Hence, a key objective in rehabilitating walking and balance is promoting the reactivation, engagement, and integration of these systems through strategies such as task-specific training, dual-tasking, strengthening, balance, and coordination training at sufficient doses, frequency, and intensity.^6,7^ However, systemic barriers such as limited therapeutic resources and inadequate payor coverage limit access to achieving the quality of rehabilitation needed to provide the greatest potential for meaningful recovery.^8,9^ To counter these real-world limitations, time-efficient, comprehensive, and effective strategies are needed to optimize post-stroke walking and balance rehabilitation and outcomes.

Backward locomotor treadmill training (BLTT) has several qualities that make it a promising strategy for comprehensive, time-efficient walking and balance rehabilitation.^10^ Since training is performed with limited body-weight support (one hand on the handrail), trainees must bear more weight on both the paretic and non-paretic lower extremities, which may improve lower limb strengthening.^11^ Additionally, backward walking has been suggested to activate key stability muscles such as the trunk, hip, and knee muscles ^12,13^, and may target the maladaptive flexor-synergy gait pattern^14,15^ to a greater extent than forward training. Since participants cannot see where they are stepping in space during BLTT, this approach may also enhance sensorineural integration by requiring greater engagement from proprioceptive and vestibular centers during training.^16,17^ For example, a recent pilot study from our group found that BLTT is safe and feasible and resulted in clinically meaningful improvement in walking speed in chronic stroke survivors following six training sessions, with retention up to two weeks post-training^10^. Despite its promising findings, the study lacked a forward-walking control group and would have benefitted from longer-term follow-up data—limiting the conclusions that could be drawn regarding the efficacy and longitudinal effects of BLTT.

Therefore, the primary objective of this two-arm randomized clinical trial was to test the preliminary efficacy of BLTT compared to forward locomotor treadmill training [(FLTT)/Control] on walking speed. Secondarily, we aimed to determine the impact of training on walking spatiotemporal symmetry, and postural stability as a surrogate measure of balance during quiet stance, as well as exploring the retention of those changes at 30- and 90 days post-training. We hypothesized that BLTT would improve walking speed, spatiotemporal symmetry, and postural stability compared to the control group and that these effects would extend beyond the training period.

## METHODS

### Participants

This study was conducted at the University of Cincinnati Neurorecovery Lab from September 2020 to January 2023. Forty community-dwelling stroke chronic stroke survivors (≥6 months post-stroke), 18-80 years of age, with mild-moderate residual walking impairment secondary to ischemic or hemorrhagic stroke, were recruited. According to the Declaration of Helsinki recommendations, all participants provided written informed consent before enrollment. As part of the institutional regular health and safety precautions during the COVID-19 pandemic, all participants were required to wear a surgical mask during training and outcome testing.

### Study Design

This study was approved by the University of Cincinnati Institutional Board Review and preregistered (clinical trial registration number: NCT04553198). Screening consisted of a history and physical exam, an overground 10-meter walk test, and a practice walk on the treadmill (3 minutes forward/ 3 minutes backward). Participants were eligible if they were: 1)18-80 years of age; 2) at least 6 months out from their stroke; 3) capable of providing consent (Mini-Mental State Exam Score >23); 4) ability to maintain ≥ 0.3mph speed (forward and backward) for a 6-minute interval on the treadmill; 5) able to walk (cane and hemi-walker acceptable) independently > 10 meters overground with the Free Step Harness System (as a safety precaution); 6) discharged from formal rehabilitation. Exclusion: 1) unstable cardiac status, which would preclude participation in a moderate intensity exercise program; 2) significant language barrier, which might prevent the participant from following instructions during training and testing; 3) adverse health condition that might affect walking capacity (severe arthritis, significant pulmonary disease significant ataxia or neglect); 4) severe lower extremity spasticity (Ashworth >2); 5) Depression (>10 on the Patient Health Questionnaire-9, if untreated). Additionally, participants were asked to abstain from formal physiotherapy and botulinum toxin treatments^18^ at least two weeks before enrollment and for the training and primary follow-up (24-hour POST). Participants meeting the inclusion and exclusion criteria were block-randomized (REDCap algorithm generated by lead statistician, H.J.S.) to either BLTT or FLTT based on self-selected overground walking speed (0.4-0.8 m/s vs. >0.8 m/s) obtained at the start of Training Day #1. Participants then underwent nine 30-minute sessions of BLTT or FLTT over three weeks, Figure 1. Trained physical therapists with experience in walking and balance rehabilitation research trials served as blinded outcome assessors.

**Figure 1.**
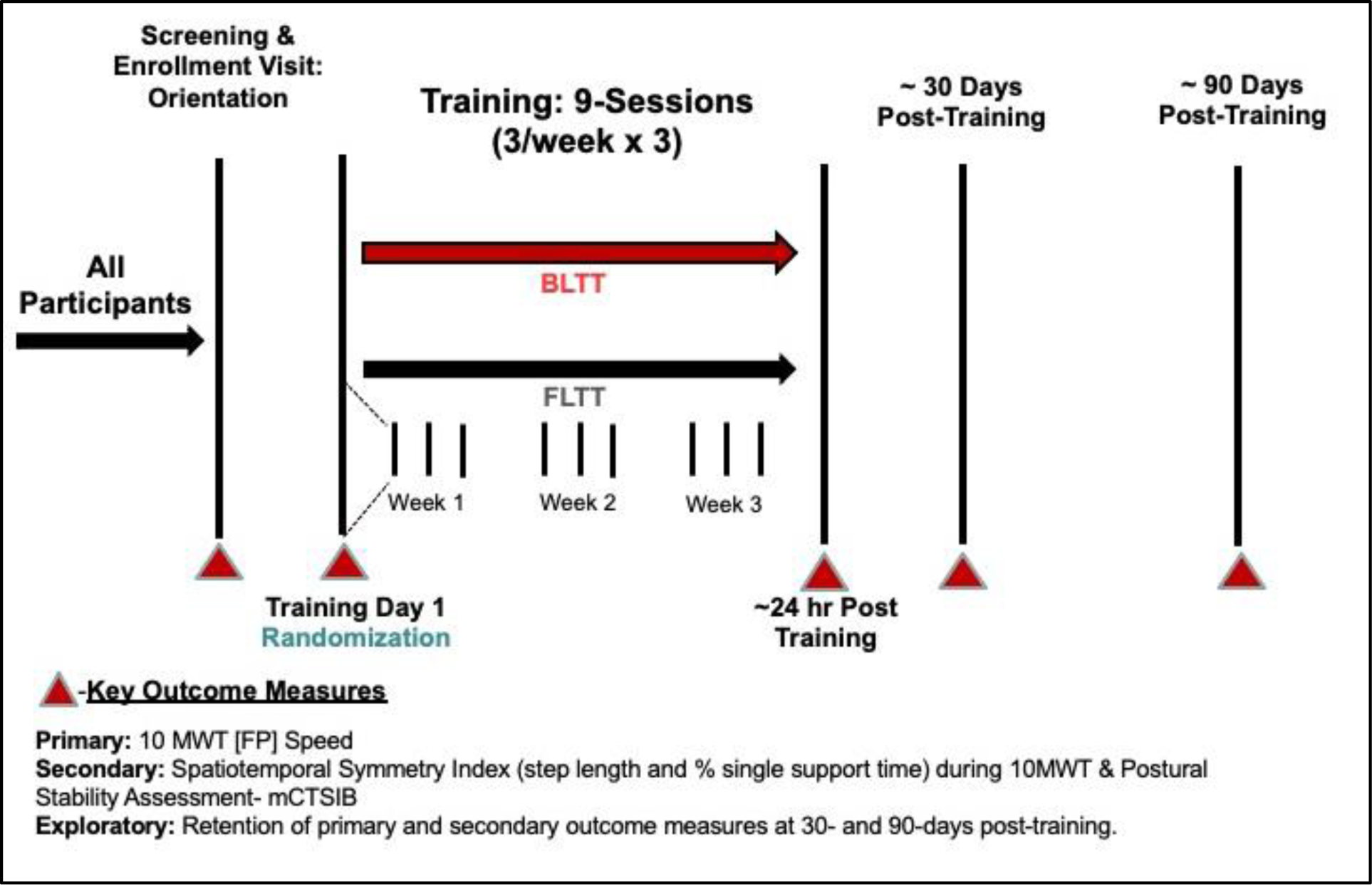
Study Schematic: 10 MWT (10-meter walk Test); FP (Fast-Paced); mCTSIB-modified Clinical test of Sensory Interaction in Balance; BLTT (backward locomotion treadmill training); FLTT (forward locomotion treadmill training

### Training protocol

As a safety precaution, participants were connected to a safety harness without body-weight support during training. Additionally, participants were required to manually hold on to the ipsilesional (less affected side) treadmill handrail continuously during training to provide standardization across sessions and between groups. The treadmill belt speed started at the most comfortable speed established during the screening visit. The speed was adjustable throughout the training period based on patient preference or per therapist recommendation, barring safety concerns. Per protocol, two-minute rest breaks were given after each bout of six minutes of treadmill training (e.g., 6min-2-min-6min-2min-6min-2min-6min). On subsequent training days (#2-9), the starting treadmill speed was set to the last treadmill speed reached on the preceding training day. To facilitate translation to overground walking, a 5-minute overground forward walking cooldown period followed each training session for both groups.

To monitor training intensity, average walking speed, steps taken, and distance traveled per day were collected—Supplementary Table 1. Additionally, to capture potential mechanistic differences between BLTT and FLTT, training-related measures, including the proportion of time on each limb (paretic/non-paretic), step length, and step length variability per session were assessed using embedded sensors on the instrumented Biodex Gait Trainer™ 3 motorized treadmill. Supplementary Figures 1-2.

### Outcome Measures

#### Overground Walking Speed

Commonly called the sixth vital sign,^19^, the 10-meter walk test (10 MWT) is the gold standard measure of post-stroke walking function that reflects overall mobility^20,21^ and health status.^22^ In this study, two 10 MWT fast trials (using a stopwatch), at fastest pace (FP) were averaged and documented in meters/second. Time points were Training Day #1 (Baseline), 24 hours, 30, and 90 days POST.

#### Spatiotemporal Symmetry During Overground Walking

Walking after stroke is often characterized by asymmetry, often leading to difficulties in balance control, falls, loss of bone mass density of the paretic hip, and increased risk of musculoskeletal pain and joint degeneration, as well as in the non-paretic limb.^23,24^ Mitigating these risks requires improvement of walking symmetry and interlimb coordination. ^25^ To test the impact of training on spatiotemporal symmetry, the average lower extremity step length (SL, spatial) and single support time (SST, temporal) symmetry were collected during the 10 MWT trials using the ProtoKinetics ZenoTM Walkway Gait Analysis System (ProtoKinetics LLC, Havertown, PA, USA), and analyzed offline by a blinded assessor. The following equation was used for measuring spatial and temporal symmetry during the 10 MWT: [1 - | Paretic – Non-paretic | / (Paretic + Non-paretic)] * 100%, where the values for the respective measure, based on the limb (i.e., paretic or nonparetic) are inputted. Possible symmetry values range from 0–100%, where 100% means perfect symmetry and 0% means complete asymmetry.^26^

#### Postural Stability

Postural stability measures sensorineural integration, motor control, and balance and is primarily mediated by visual, somatosensory, and vestibular centers. The modified clinical test of sensory interaction on balance (mCTSIB)^27-29^ is a reliable measure of postural stability and was utilized to capture the impact of training on sensory systems processing. Participants stood on a platform (with safety harness) with their hands at their side under four distinct conditions (30 secs/condition): 1) firm surface with the eyes open (testing integration of all sensory modalities), 2) firm surface with the eyes closed (testing integration between somatosensory and vestibular systems in the absence of vision), 3) compliant surface (foam) with the eyes open (testing integration of visual and vestibular pathways during perturbation of somatosensory input), 4) Compliant surface (foam) with the eyes closed (testing the integrity of the vestibular system, in the absence of visual input and perturbed somatosensory input). The output measure for each mCTSIB condition was the sway velocity index (SVI, degree of sway/secs), representing the mean velocity of the center of gravity sway during that condition, with higher values representing greater instability. Under each condition, the SVIs from three trials were collected and averaged.

#### Sample Size Determination

Given the limited number of adequately powered walking rehabilitation studies comparing backward versus forward treadmill training in literature, the study by Yang and colleagues,^30^, which tested backward walking (overground) combined with conventional physiotherapy on walking outcomes in chronic stroke survivors, was used for sample size determination. Based on their reported change in overground walking speed, this study was powered to detect a between-group difference of 0.11 m/s on the 10 MWT (fast) from baseline to 24-hour post-training. We anticipated that 20 participants in each group were sufficient to achieve an 80% power to detect this mean difference, with an estimated standard deviation of 0.12 m/s in both groups using a two-sample t-test.

#### Statistical Analyses

Statistical analyses followed a prespecified plan, used an intent-to-treat approach, and were done with SAS, version 9.4 (SAS Institute Inc). The primary study statistician (H.J.S.) remained blinded to treatment groups until after the primary analysis. Normality assumption was tested by the Shapiro-Wilk method, and the significance level was set at p=0.05 for all measures. Linear mixed-effects models were used to test for within-group change and between-group differences stability (Δ=24hr Post Training-Training Day 1) in change for walking speed, spatiotemporal symmetry index, and postural stability. These models included each walking measure (separately) as the dependent variable, with fixed effects for time point, group, and their interaction and a random effect for participants to account for the correlated nature of repeated measures from the same person. The retention of effect within and between groups on each walking measure was determined by comparing the change between 30- and 90-days POST and baseline.

## RESULTS

In total, 44 chronic stroke survivors were screened, and 40 were randomized to either BLTT (n=19) or FLTT (n=21), equating to a 90% enrollment rate, Figure 2. The mean age of enrollees was 56.3 ± 8.6 years; 53% were female, and 30% were non-Hispanic Black, Table 1. There were no significant differences between groups regarding baseline demographics and metrics, except for survivors using a single-point cane which was all in the FLTT group.

**Figure 2.**
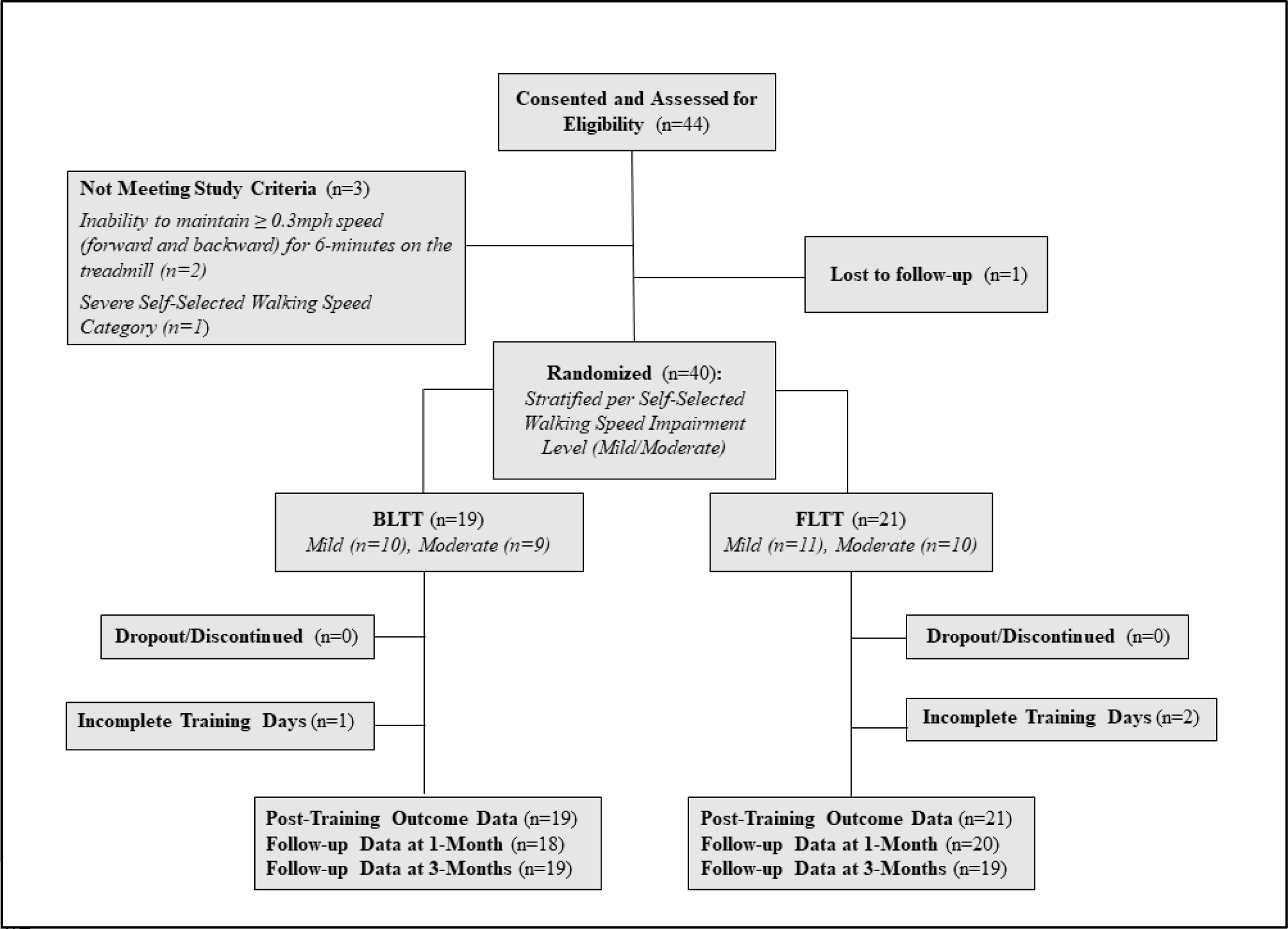
Trial Profile

**Table 1.**
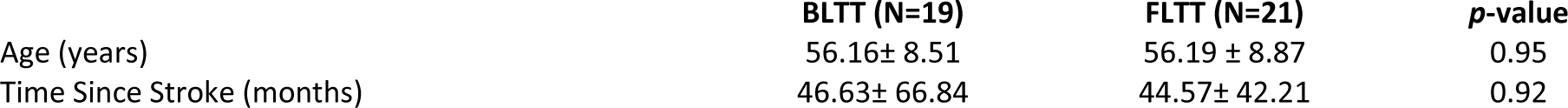
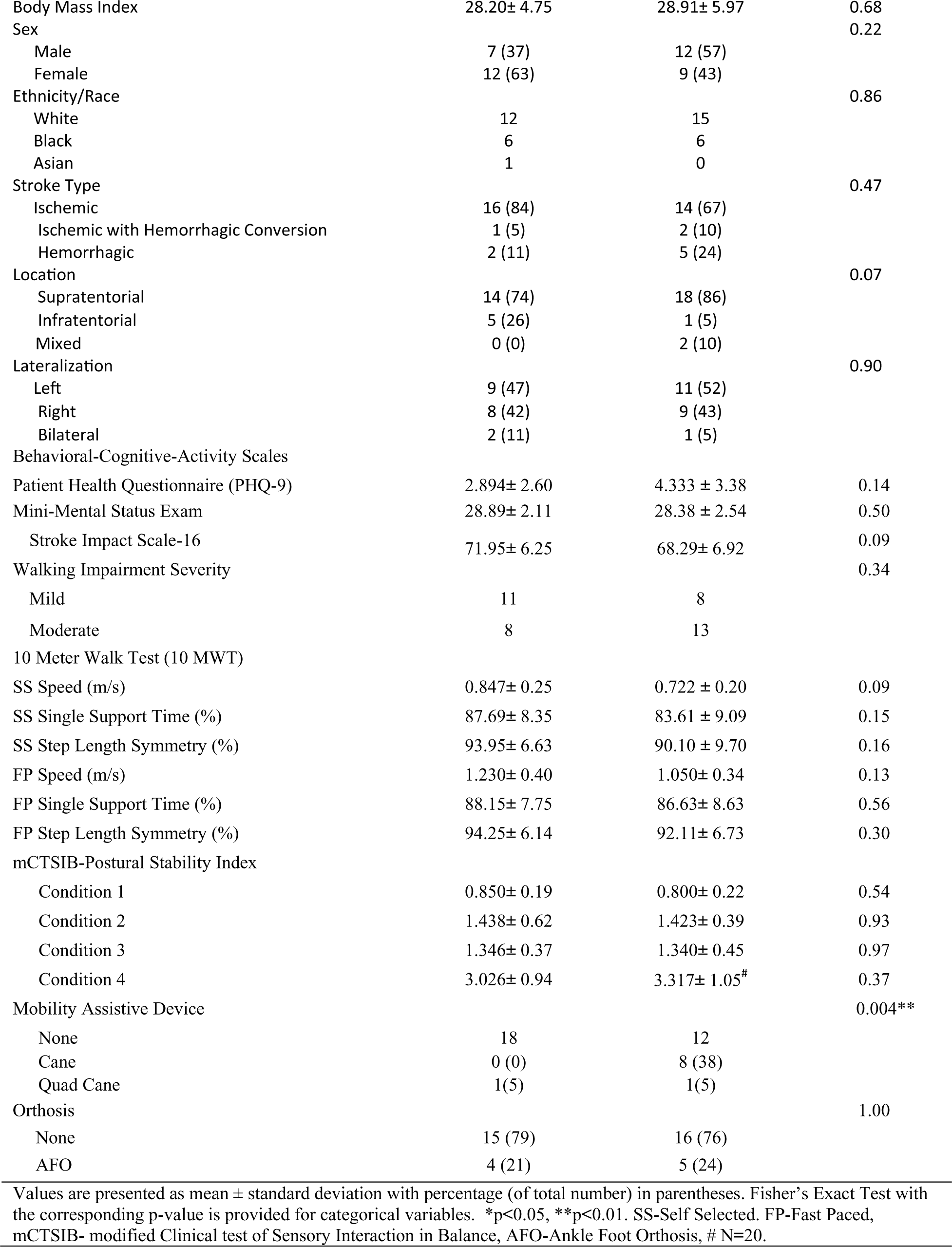
Baseline Demographic, Gait, and Postural Stability Variables per Training Group.

|  | **BLTT (N=19)** | **FLTT (N=21)** | ***p*-value** |
| --- | --- | --- | --- |
| Age (years) | 56.16± 8.51 | 56.19 ± 8.87 | 0.95 |
| Time Since Stroke (months) | 46.63± 66.84 | 44.57± 42.21 | 0.92 |
| Body Mass Index | 28.20± 4.75 | 28.91± 5.97 | 0.68 |
| Sex |  |  | 0.22 |
| Male | 7 (37) | 12 (57) |  |
| Female | 12 (63) | 9 (43) |  |
| Ethnicity/Race |  |  | 0.86 |
| White | 12 | 15 |  |
| Black | 6 | 6 |  |
| Asian | 1 | 0 |  |
| Stroke Type |  |  | 0.47 |
| Ischemic | 16 (84) | 14 (67) |  |
| Ischemic with Hemorrhagic Conversion | 1 (5) | 2 (10) |  |
| Hemorrhagic | 2 (11) | 5 (24) |  |
| Location |  |  | 0.07 |
| Supratentorial | 14 (74) | 18 (86) |  |
| Infratentorial | 5 (26) | 1 (5) |  |
| Mixed | 0 (0) | 2 (10) |  |
| Lateralization |  |  | 0.90 |
| Left | 9 (47) | 11 (52) |  |
| Right | 8 (42) | 9 (43) |  |
| Bilateral | 2 (11) | 1 (5) |  |
| Behavioral-Cognitive-Activity Scales |  |  |  |
| Patient Health Questionnaire (PHQ-9) | 2.894± 2.60 | 4.333 ± 3.38 | 0.14 |
| Mini-Mental Status Exam | 28.89± 2.11 | 28.38 ± 2.54 | 0.50 |
| Stroke Impact Scale-16 | 71.95± 6.25 | 68.29± 6.92 | 0.09 |
| Walking Impairment Severity |  |  | 0.34 |
| Mild | 11 | 8 |  |
| Moderate | 8 | 13 |  |
| 10 Meter Walk Test (10 MWT) |  |  |  |
| SS Speed (m/s) | 0.847± 0.25 | 0.722 ± 0.20 | 0.09 |
| SS Single Support Time (%) | 87.69± 8.35 | 83.61 ± 9.09 | 0.15 |
| SS Step Length Symmetry (%) | 93.95± 6.63 | 90.10 ± 9.70 | 0.16 |
| FP Speed (m/s) | 1.230± 0.40 | 1.050± 0.34 | 0.13 |
| FP Single Support Time (%) | 88.15± 7.75 | 86.63± 8.63 | 0.56 |
| FP Step Length Symmetry (%) | 94.25± 6.14 | 92.11± 6.73 | 0.30 |
| mCTSIB-Postural Stability Index |  |  |  |
| Condition 1 | 0.850± 0.19 | 0.800± 0.22 | 0.54 |
| Condition 2 | 1.438± 0.62 | 1.423± 0.39 | 0.93 |
| Condition 3 | 1.346± 0.37 | 1.340± 0.45 | 0.97 |
| Condition 4 | 3.026± 0.94 | 3.317± 1.05<sup>#</sup> | 0.37 |
| Mobility Assistive Device |  |  | 0.004** |
| None | 18 | 12 |  |
| Cane | 0 (0) | 8 (38) |  |
| Quad Cane | 1(5) | 1(5) |  |
| Orthosis |  |  | 1.00 |
| None | 15 (79) | 16 (76) |  |
| AFO | 4 (21) | 5 (24) |  |

Values are presented as mean ± standard deviation with percentage (of total number) in parentheses. Fisher's Exact Test with the corresponding p-value is provided for categorical variables. \*p<0.05, \*\*p<0.01. SS-Self Selected. FP-Fast Paced, mCTSIB- modified Clinical test of Sensory Interaction in Balance, AFO-Ankle Foot Orthosis, # N=20.

### Training, Safety, and Tolerability

Ninety-eight percent (167/171) of BLTT and ninety-seven percent (183/189) of FLTT sessions were completed. The mean change in treadmill training speed from baseline to the end of training was 0.27 m/s ±0.14 for BLTT and 0.29 m/s ±0.14 for FLTT. The mean total step count achieved over nine sessions training was 12,756 ±3,758 for BLTT and 16,403 ±4,371 for FLTT, see supplementary Table 1. The BLTT group spent more time on the paretic than the non-paretic leg relative to FLTT, Supplementary Figure 1. Additionally, participants in the BLTT group demonstrated improvement in bilateral step length variabilities relative to baseline, whereas participants in FLTT overall demonstrated lower but unchanged variability, Supplementary Figure 2. No serious adverse events were reported in either group, with the most common complaint being transient soreness (BLTT-63%, FLTT-48%) and fatigue (BLTT-63%, FLTT-52%) related to training, Supplementary Table 2. Additionally, both groups experienced comparable improvement in activity level, perceived strength, energy level, and mood, Supplementary Table 3.

### Follow-up

All randomized participants reached the primary endpoint (24-hr POST). For the 30- and 90-day follow-ups, one participant in the BLTT group missed Day 30 POST due to illness with COVID-19 but was able to return for the Day 90 POST. Two participants from the FLTT group were lost to follow-up: the first after 24-hr POST and the second following Day 30 POST.

### Fast Walking Speed (Primary Outcome)

Both BLTT and FLTT groups experienced an improvement in FP walking speed (p<0.0001) with no between-group differences (p=0.26). The mean change in walking speed was 0.22 m/s (0.14-0.29) for BLTT and 0.24 m/s (0.17-0.31) for FLTT. Retention of speed relative to baseline was 0.13 m/s (0.06-0.20) for BLTT and 0.19 m/s (0.12-0.26) for FLTT at 30 days post-training, with no between-group differences (p=0.34). At day 90, retention was 0.18 m/s (0.11-0.24) for BLTT and 0.19 (0.12-0.26) for FLTT, respectively, with no between-group differences (p=0.79), Figure 3.

**Figure 3.**
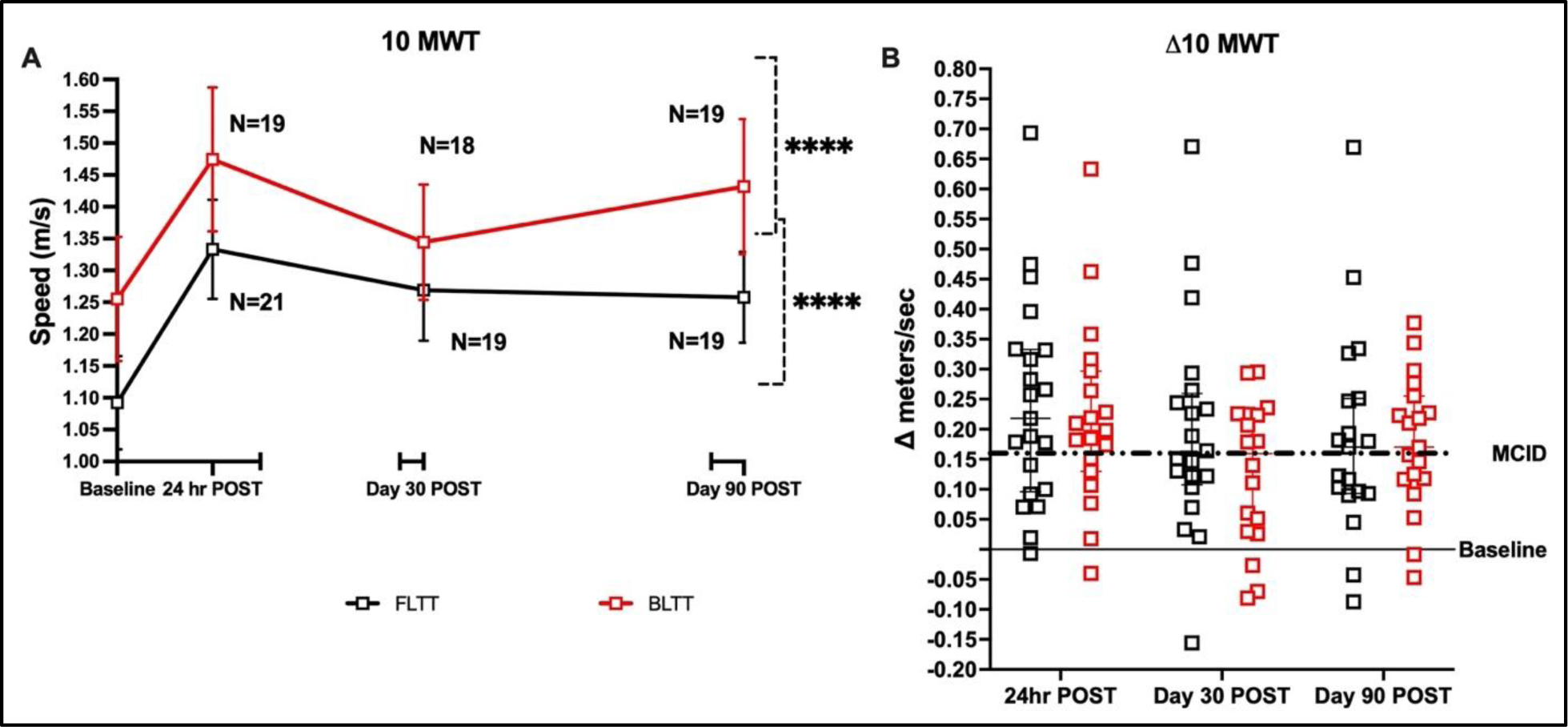
Fast Walking Speed. Mean walking 10-meter walk test (10MWT) speed pre-90 days post-training (A), represented as the mean and standard error measure. Retention of walking speed post-training, 30- and 90-days post-training, with 0.16 m/s (dotted line) as references for the minimal clinically important difference (MCID)^20^, (B). Squares represent individual participant data at each time point. ****p<0.0001

### Spatiotemporal Symmetry Index

#### Step Length Symmetry (SLS)

No significant improvement in SLS was seen within (p=0.12) or between groups (p= 0.20) immediately post-training, Figure 4A. The mean change in SLS post-training was 0.59 (-1.39, 2.58) for BLTT and 0.55 (: -0.54, 3.22) for FLTT. On 30-day POST, the BLTT group showed within-group improvement in SLS change relative to baseline, 1.96 (0.17, 3.76), p=0.03, unlike the FLTT group, 0.03 (-0.92, 2.51), p=0.35; however, no between-group difference was seen, p= 0.35. On 90-day POST, there was slight regression in SLS compared to 30-day POST in the BLTT [ 1.37 (-0.25, 2.98), p=0.09] and FLTT [ 0.01 (-1.48, 1.68), p=0.90], with no between-group difference, p=0.26. See Table 2.

**Figure 4.**
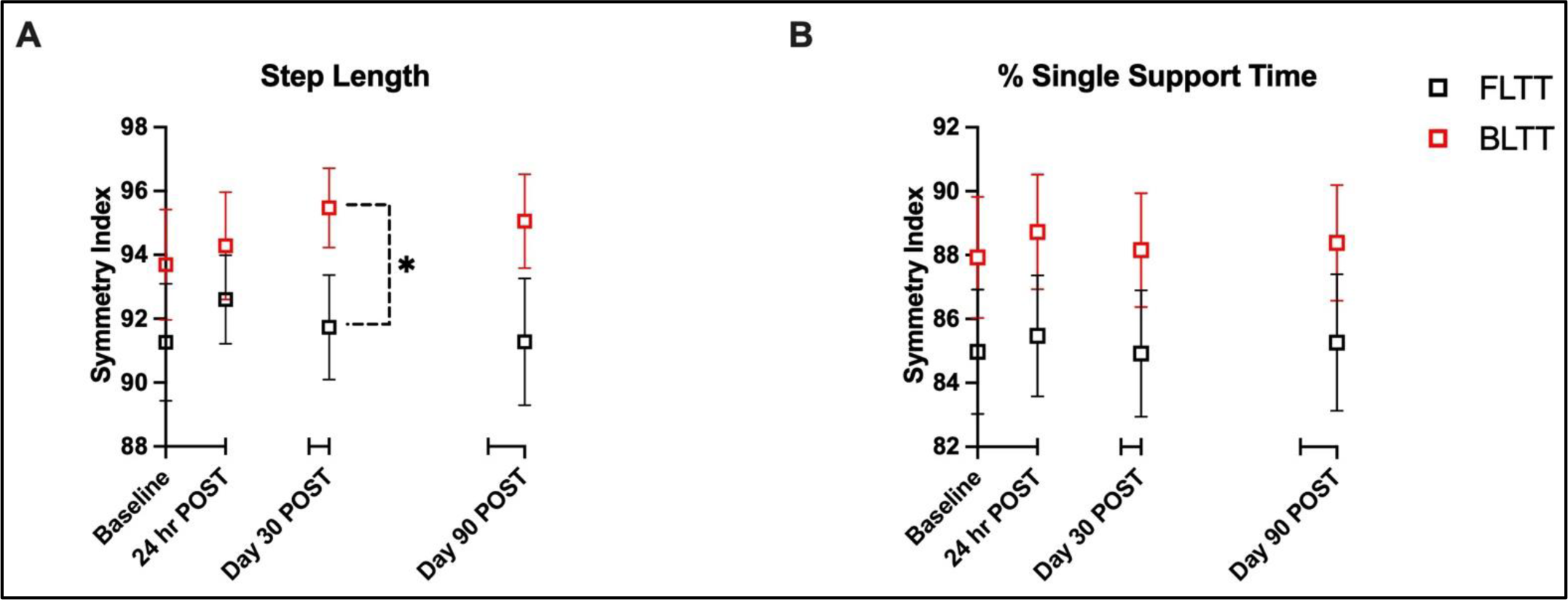
Spatiotemporal Symmetry Index. Step Length Symmetry(A) % Single Support Time Symmetry (B) represents the mean and standard error measure. *p<0.05.

#### % Single Support Time Symmetry (% SSTS)

No significant improvement in % SSTS was seen within (p=0.62) or between groups (p= 0.25) immediately post-training, Figure 4B. The mean change in % SSTS was 0.79 (-0.11, 0.08) for BLTT and 0.50 (-1.29, 2.30) for FLTT. No within- or between-group differences were seen at 30-day POST and 90-day POST.

#### Postural Stability

*Condition #1*: All participants completed Condition #1, testing the quality of integration of visual, somatosensory, and vestibular toward postural stability. No significant improvement (decrease) in postural stability (SVI) scores was seen within (p=0.77) or between groups (p=0.96) post-training. The mean change in SVI post-training was -0.01 (-0.11, 0.08) for BLTT and -0.02 (-0.11, 0.08) for FLTT. On Day 30, the retention of SVI was minimally improved at -0.07 (-0.21, 0.07) for BLTT and 0.00 (-0.13, 0.14) FLTT, with no between-group differences (p=0.46). At day 90 POST, compared to day 30 POST retention was sustained for BLTT [-0.10 (-0.22, 0.02)] and regressed for FLTT [0.04 (-0.08, 0.15)]; however, no with or between-group differences were appreciated (p=0.10) Figure 5a.

**Figure 5.**
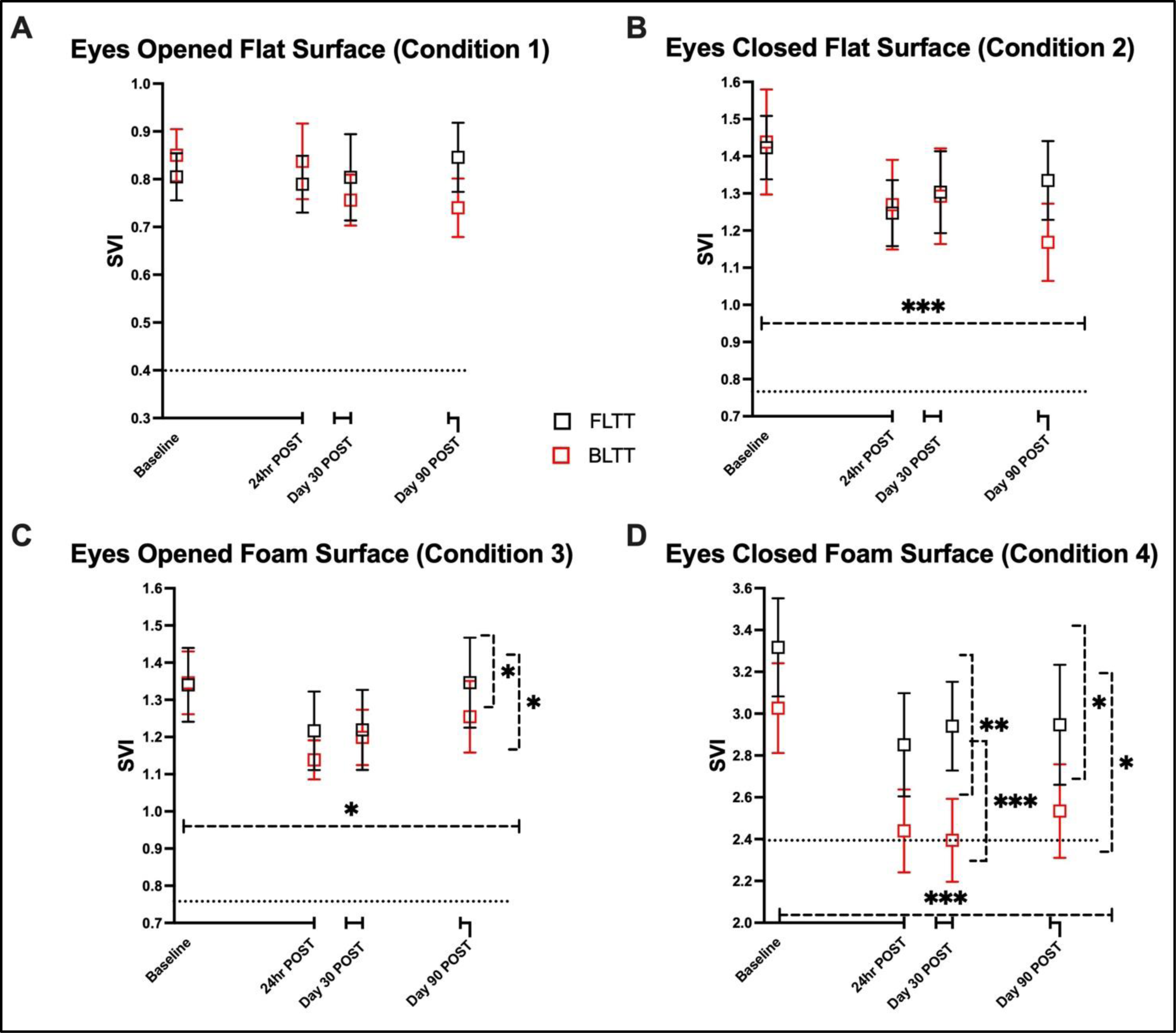
Sway Velocity Index [SVI) on the modified Clinical test of Sensory Interaction in Balance of the Balance Platform Biodex Balance System™, represented as the mean and standard error measure. The dotted lines represent normative data for each condition.^31^ *p<0.05, **p<0.01, ***p<0.001, ****p<0.0001

*Condition #2:* All participants completed Condition #2, testing the interaction of somatosensory and vestibular systems on postural stability in the absence of vision. Both groups experienced improvement in SVI with training (p=0.01). The mean change in SVI post-training was [-0.17 (- 0.33, -0.01), p=0.04] for BLTT and [ -0.18 (-0.33, -0.02), p=0.03] for FLTT, with no between-group difference, p=0.95). On Day 30 POST, the retention of SVI decreased to [ -0.09 (-0.23, 0.04), p=0.18) for BLTT and sustained at [-0.14 (-0.27, -0.01), p=0.03] for FLTT, with no between-group difference (p=0.60). On day 90 POST, retention was improved for BLTT [ -0.20 (-0.33, -0.06), p=0.005) and reduced for FLTT [-0.12 (-0.25, 0.01), p=0.07)]; however, no between-group differences were appreciated (p=0.42) Figure 5B.

*Condition #3:* All participants completed Condition #3, testing visual and vestibular contributions to postural stability. Both groups experienced improvement in SVI with training (p=0.04). The mean change in SVI post-training was [-0.21 (-0.36, -0.05), p=0.01] for BLTT and [ -0.12 (-0.27, 0.02), p=0.09] for FLTT, with no between-group difference, p=0.43). On Day 30 POST, the retention of SVI decreased to [ -0.13 (-0.26, 0.01), p=0.06) for BLTT and sustained for FLTT [-0.14 (-0.27, -0.01), p=0.03], with no between-group difference (p=0.86). On day 90 POST, retention was reduced for BLTT [ -0.09 ( -0.31,0.13), p=0.41) and FLTT [-0.02 (-0.23, 0.19), p=0.83)], with no between-group differences (p=0.66) Figure 5C.

*Condition #4:* Ninety-eight percent (39/40) of participants completed Condition #4 at baseline, testing vestibular contributions to postural stability. One hundred percent completed Condition #4 at 24hr POST. Overall, both groups experienced an improvement in SVI with training (p<0.0002). The mean change in SVI post-training was [-0.59 (-0.88, -0.29), p=0.0003] for BLTT and [ -0.48 (-0.77, -0.20), p=0.002] for FLTT, with no between-group difference, p=0.61). On Day 30 POST, the retention of SVI was sustained at [ -0.57 (-0.85, -0.28), p=0.0003) for BLTT and [-0.43 (-0.71, -0.15), p=0.004] for FLTT, with no between-group difference (p=0.49). On day 90 POST, retention remained sustained for BLTT [ -0.48( -0.87, -0.09), p=0.02) and FLTT [-0.44 (-0.82, -0.05), p=0.03)], with no between-group differences (p=0.87) Figure 5D.

## DISCUSSION

The primary objective of this study was to test the preliminary efficacy of BLTT compared to FLTT on overground walking speed in chronic stroke survivors with mild-moderate walking impairment. It also aimed to characterize the long-term effects of training on walking and postural stability measures. As anticipated, BLTT resulted in long-lasting and clinically meaningful improvement in overground walking speed up to 90 days post-training. However, an unexpected finding was that FLTT resulted similarly in a significant improvement in walking speed. Although this study was not powered to detect significance for symmetry and postural stability measures, we found that BLTT resulted in greater offline improvement in step length symmetry and point-estimates suggested the potential for longer-lasting improvements in subcomponents of the mCTSIB-- possibly driven by proprioceptive and vestibular integration (i.e., Conditions 1 and 2).

### Mechanistic Differences in Training

This study found that both the backward and forward training groups experienced a sizable and long-lasting improvement in forward walking speed following just nine training sessions (∼270 minutes). This favorable improvement may have resulted from the rigor and intensity of our training approach, which required participants to support much of their body weight while keeping up with and maintaining their balance on a constantly moving treadmill. Additionally, participants were able to perform thousands of repetitions per training day, compared to conventional physiotherapy, where fewer repetitions are achieved.^32^ Indeed, future investigations are warranted to determine if a longer duration of training, beyond nine sessions, can compound the improvements observed in this study.

The inclusion of both a backward and forward walking group provided an opportunity to assess over time how backward may differentially impact walking and postural stability.^13^ While both groups experienced improvement in their walking speed, the mechanism responsible for these training-related effects may differ. For example, consistent with observations from past walking rehabilitation studies and clinical practice guidelines, the improvement in walking speed in the FLTT may have been a result of the task-specific nature of this training approach, where participants were able to achieve higher training speeds and repetitions, translating to faster overground walking speeds.^33^ Nevertheless, despite the improvement in walking speed, there was no notable improvement in walking symmetry in the FLTT group—suggesting that the improvement in walking speed could be related to adaptation and the development of compensatory strategies and less so motor control and plasticity. ^34^ In contrast, while participants in the BLTT, on average, experienced fewer walking repetitions and trained at slower treadmill speeds (Supplementary Table 1) the observed change in overground walking speed was comparable to FLTT. Moreover, the BLTT group demonstrated progressive improvement and retention of step length symmetry and postural stability, particularly in the absence of vision (Condition #2). While the reason for this finding is multifactorial, one possibility is that BLTT better targets motor control, promoting longer stance duration on the paretic limb (see Supplementary Figure 1) while concurrently training sensorineural integration through greater reliance on proprioceptive and vestibular senses to maintain posture during training.^17,35,36^ Therefore, the improvement and retention of step length symmetry and postural stability, particularly in the absence of vision (Condition #2), may have been due to the culmination of strength and sensorineural system training experienced with BLTT, suggesting that this training approach may better improve walking mechanics and promote better postural stability at higher walking speeds. Notably, the observation that these changes were more pronounced offline suggests that these training-associated changes are not immediate and may require time (i.e., days to weeks) to translate to improvements in overground walking patterns.^37^

Our working hypothesis that BLTT better targets the paretic limb contrasts with a recent study in chronic stroke survivors testing overground backward versus forward walking training,^38^ which found that overground backward training did not improve the propulsive force generated by the paretic limb and instead improved the propulsion of the non-paretic limb. The contrasting outcome may be related to the inherent differences in treadmill versus overground training. Specifically, akin to studies using split-belt treadmill training,^39,40^ The prolonged stance phase during the BLTT protocol is likely due to the additional time and effort it takes for subjects to activate the paretic limb to the extent of generating enough force during single-limb support to enable the successful backward extension (swing) of the paretic limb. Over time, this pattern may lead to greater strengthening of the paretic limb and is more likely to result in higher paretic propulsion.^41,42^ Another possibility, from a temporal muscle activation standpoint,^12^ is that BLTT was able to target better the maladaptive flexor-synergy gait pattern associated with stroke through forced hip extension and ankle dorsiflexion of the paretic limb.^17,43^

In contrast our findings with step length symmetry, BLTT had no significant impact on temporal symmetry. This finding is consistent with past walking rehabilitation studies,^40,44-46^ suggesting that the modulation of percent single support time symmetry with treadmill training can be challenging, since training on a constantly moving platform often results in the same stance time patterns, with limited opportunities for stride-by-stride adjustments. Therefore, it is likely that additional training, beyond nine sessions, is needed to observe improvements in this measure.

### Vestibular Processing and Post-Stroke Walking and Balance Recovery

Vestibular hypofunction is a relatively underdiagnosed consequence of stroke^47^ and may be related to injury to infra and supratentorial vestibular centers or may be a result of post-stroke inactivity and deconditioning.^26^ In this study, both groups experienced long-lasting improvements in a marker of vestibular processing after nine training sessions, see Figure 5D. Although we observed no significant difference between groups, the magnitude of improvement seen post-training was nearly three-fold greater compared to the other conditions--with the BLTT cohort achieving normative levels at Day 30 post-training. While the vestibular system represents just one subsection of sensory system processing and upstream motor control, it may play a more significant role in post-stroke walking and balance recovery than classically thought. Indeed, the full extent to which our training approach can improve vestibular function and other aspects of sensory systems remains undetermined and warrants further investigation in future higher-dosed and well-powered walking and balance rehabilitation studies.

### Limitations

While careful attention was taken in designing the randomization process, we found that a higher proportion of participants in the FLTT group used a cane at baseline. This observation resulted from chance and did not appear to have any bearing on other baseline measures, such as walking speed, symmetry, or postural stability. Additionally, secondary analysis removing those participants using a cane at baseline did not significantly impact the study results. Moreover, the postulation that BLTT may have improved paretic limb strength was based on deductive reasoning and not empirically measured in this study. Therefore, future mechanistic and appropriately powered studies should incorporate ground reaction forces and kinematics to test this working hypothesis. Lastly, stroke survivors with severe walking impairment and non-ambulators were not included in this study, limiting the generalizability of our findings.

## CONCLUSION

BLTT resulted in clinically meaningful changes in walking speed after nine training sessions, with sustained effects up to 90 days post-training; however, no differences were observed between groups (BLTT vs FLTT). Additionally, BLTT improved spatial symmetry, most notably at 30 days post-training, and resulted in offline enhancement of proprio-vestibular processing, leading toward a significant improvement in postural stability. Therefore, BLTT may be a useful and comprehensive rehabilitation strategy for optimizing walking and balance outcomes.

## Data Availability

Deidentified data is available upon request.

## ACKNOWLEDGMENTS

The authors thank the study participants for their time and effort.

## CITATIONS

1. Verma R, Arya K, Sharma P, Garg RK. Understanding gait control in post-stroke: Implications for management. J Bodywork Movement Ther. 2012;16(1):14–21. doi: 10.1016/j.jbmt.2010.12.005.

2. Schmid AA, Van Puymbroeck M, Altenburger PA, et al. Balance and balance self-efficacy are associated with activity and participation after stroke: A cross-sectional study in people with chronic stroke. Arch Phys Med Rehabil. 2012;93(6):1101–1107. doi: 10.1016/j.apmr.2012.01.020.

3. Moore SA, Boyne P, Fulk G, Verheyden G, Fini NA. Walk the talk: Current evidence for walking recovery after stroke, future pathways and a mission for research and clinical practice. Stroke. 2022;53(11):3494–3505. https://pubmed.ncbi.nlm.nih.gov/36069185/. Accessed Jul 29, 2024. doi: 10.1161/STROKEAHA.122.038956.

4. Van Criekinge T, Heremans C, Burridge J, et al. Standardized measurement of balance and mobility post-stroke: Consensus-based core recommendations from the third stroke recovery and rehabilitation roundtable. Int J Stroke. 2024;19(2):158–168. Accessed Feb 14, 2024. doi: 10.1177/17474930231205207.

5. Takakusaki K. Neurophysiology of gait: From the spinal cord to the frontal lobe. Movement Disorders. 2013;28(11):1483–1491. doi: 10.1002/mds.25669.

6. Langhorne P, Bernhardt J, Kwakkel G. Stroke rehabilitation. Lancet. 2011;377(9778):1693–1702. https://pubmed.ncbi.nlm.nih.gov/21571152/. Accessed Jul 29, 2024. doi: 10.1016/S0140-6736(11)60325-5.

7. Kwakkel G, Stinear C, Essers B, et al. Motor rehabilitation after stroke: European stroke organisation (ESO) consensus-based definition and guiding framework. Eur Stroke J. 2023;8(4):880–894. https://pubmed.ncbi.nlm.nih.gov/37548025/. Accessed Jul 29, 2024. doi: 10.1177/23969873231191304.

8. Egan MY, Kessler D, Ceci C, et al. Problematising risk in stroke rehabilitation. Disabil Rehabil. 2016;38(23):2334–2344. https://pubmed.ncbi.nlm.nih.gov/26731429/. Accessed Jul 29, 2024. doi: 10.3109/09638288.2015.1123304.

9. Chavez AA, Simmonds KP, Venkatachalam AM, Ifejika NL. Health care disparities in stroke rehabilitation. Phys Med Rehabil Clin N Am. 2024;35(2):293–303. https://pubmed.ncbi.nlm.nih.gov/38514219/. Accessed Jul 29, 2024. doi: 10.1016/j.pmr.2023.06.030.

10. Awosika OO, Chan D, Sucharew HJ, et al. Backward locomotor treadmill training differentially improves walking performance across stroke walking impairment levels. Brain Sci. 2022;12(2):133. Accessed May 30, 2023. doi: 10.3390/brainsci12020133.

11. Wernig A, Wernig S. The trouble with “Body weight support” in treadmill training. Arch Phys Med Rehabil. 2010;91(9):1478. doi: 10.1016/j.apmr.2010.05.015.

12. Winter DA, Pluck N, Yang JF. Backward walking: A simple reversal of forward walking? J Mot Behav. 1989;21(3):291–305. doi: 10.1080/00222895.1989.10735483.

13. Thorstensson A. How is the normal locomotor program modified to produce backward walking? Experimental Brain Research. 1986;61(3):664–668. doi: 10.1007/BF00237595.

14. Schneider C, Capaday C. Progressive adaptation of the soleus H-reflex with daily training at walking backward. J Neurophysiol. 2003;89(2):648–656. doi: 10.1152/jn.00403.2002.

15. Winter DA. Biomechanics of normal and pathological gait: Implications for understanding human locomotor control. J Mot Behav. 1989;21(4):337–355.

16. Fritz NE, Worstell AM, Kloos AD, Siles AB, White SE, Kegelmeyer DA. Backward walking measures are sensitive to age-related changes in mobility and balance. Gait \& Posture. 2013;37(4):593–597. doi: 10.1016/j.gaitpost.2012.09.022.

17. Rose DK, DeMark L, Fox EJ, Clark DJ, Wludyka P. A backward walking training program to improve balance and mobility in acute stroke. Journal of Neurologic Physical Therapy. 2018;42(1):12–21. doi: 10.1097/NPT.0000000000000210.

18. Jacinto J, Varriale P, Pain E, Lysandropoulos A, Esquenazi A. Patient perspectives on the therapeutic profile of botulinum neurotoxin type A in spasticity. Frontiers in Neurology. 2020;11:388. doi: 10.3389/fneur.2020.00388.

19. Middleton A, Fritz SL, Lusardi M. Walking speed: The functional vital sign. J Aging Phys Act. 2015;23(2):314–322. Accessed May 30, 2023. doi: 10.1123/japa.2013-0236.

20. Schmid A, Duncan PW, Studenski S, et al. Improvements in speed-based gait classifications are meaningful. Stroke. 2007;38(7):2096–2100. doi: 10.1161/STROKEAHA.106.475921.

21. Lord SE, McPherson K, McNaughton HK, Rochester L, Weatherall M. Community ambulation after stroke: How important and obtainable is it and what measures appear predictive?1 1No commercial party having a direct financial interest in the results of the research supporting this article has or will confer a benefit on the author(s) or on any organization with which the author(s) is/are associated. Arch Phys Med Rehabil. 2004;85(2):234–239. doi: 10.1016/j.apmr.2003.05.002.

22. Studenski S, Perera S, Wallace D, et al. Physical performance measures in the clinical setting. J Am Geriatr Soc. 2003;51(3):314–322. doi: 10.1046/j.1532-5415.2003.51104.x.

23. Patterson KK, Parafianowicz I, Danells CJ, et al. Gait asymmetry in community-ambulating stroke survivors. Arch Phys Med Rehabil. 2008;89(2):304–310. doi: 10.1016/j.apmr.2007.08.142.

24. Jørgensen L, Jacobsen BK. Changes in muscle mass, fat mass, and bone mineral content in the legs after stroke: A 1 year prospective study. Bone. 2001;28(6):655–659. doi: 10.1016/S8756-3282(01)00434-3.

25. Cleland BT, Schindler-Ivens S. Symmetry is associated with interlimb coordination during walking and pedaling after stroke. J Neurol Phys Ther. 2022;46(2):81–87. https://pubmed.ncbi.nlm.nih.gov/34507343/. Accessed Jul 29, 2024. doi: 10.1097/NPT.0000000000000377.

26. Awosika OO, Garver A, Drury C, et al. Insufficiencies in sensory systems reweighting is associated with walking impairment severity in chronic stroke: An observational cohort study. Front Neurol. 2023;14:1244657. doi: 10.3389/fneur.2023.1244657.

27. Antoniadou E, Kalivioti X, Stolakis K, et al. Reliability and validity of the mCTSIB dynamic platform test to assess balance in a population of older women living in the community. J Musculoskelet Neuronal Interact. 2020;20(2):185–193. https://www.ncbi.nlm.nih.gov/pmc/articles/PMC7288384/. Accessed Oct 19, 2023.

28. Peller A, Garib R, Garbe E, et al. Validity and reliability of the NIH toolbox® standing balance test as compared to the biodex balance system SD. Physiother Theory Pract. 2023;39(4):827–833. Accessed Oct 19, 2023. doi: 10.1080/09593985.2022.2027584.

29. Shumway-Cook A, Horak FB. Assessing the influence of sensory interaction of balance. suggestion from the field. Phys Ther. 1986;66(10):1548–1550.

30. Yang Y, Yen J, Wang R, Yen L, Lieu F. Gait outcomes after additional backward walking training in patients with stroke: A randomized controlled trial. Clin Rehabil. 2005;19(3):264–273. https://pubmed.ncbi.nlm.nih.gov/15859527/. Accessed Jul 29, 2024. doi: 10.1191/0269215505cr860oa.

31. Overman D. Biodex balance system SD and BioSway devices receive enhancements. Rehab Management (Online). Jun 17, 2019. Available from: https://search.proquest.com/docview/2241741150.

32. Lang CE, Macdonald JR, Reisman DS, et al. Observation of amounts of movement practice provided during stroke rehabilitation. Arch Phys Med Rehabil. 2009;90(10):1692–1698. doi: 10.1016/j.apmr.2009.04.005.

33. Hornby TG, Reisman DS, Ward IG, et al. Clinical practice guideline to improve locomotor function following chronic stroke, incomplete spinal cord injury, and brain injury. J Neurol Phys Ther. 2020;44(1):49–100. https://pubmed.ncbi.nlm.nih.gov/31834165/. Accessed Jul 29, 2024. doi: 10.1097/NPT.0000000000000303.

34. Allen JL, Kautz SA, Neptune RR. Step length asymmetry is representative of compensatory mechanisms used in post-stroke hemiparetic walking. Gait Posture. 2011;33(4):538–543. https://pubmed.ncbi.nlm.nih.gov/21316240/. Accessed Jul 29, 2024. doi: 10.1016/j.gaitpost.2011.01.004.

35. Wolpert DM, Diedrichsen J, Flanagan JR. Principles of sensorimotor learning. Nature Reviews Neuroscience. 2011;12(12):739–751. doi: 10.1038/nrn3112.

36. Hao W, Chen Y. Backward walking training improves balance in school-aged boys. *Sports Medicine, Arthroscopy, Rehabilitation*, Therapy \& Technology. 2011;3(1):24. doi: 10.1186/1758-2555-3-24.

37. Dayan E, Cohen LG. Neuroplasticity subserving motor skill learning. Neuron. 2011;72(3):443–454. doi: 10.1016/j.neuron.2011.10.008.

38. Bansal K, Vistamehr A, Conroy CL, Fox EJ, Rose DK. The influence of backward versus forward locomotor training on gait speed and balance control post-stroke: Recovery or compensation? J Biomech. 2023;155:111644. https://pubmed.ncbi.nlm.nih.gov/37229888/. Accessed Jul 29, 2024. doi: 10.1016/j.jbiomech.2023.111644.

39. Awad LN, Binder-Macleod SA, Pohlig RT, Reisman DS. Paretic propulsion and trailing limb angle are key determinants of long-distance walking function after stroke. Neurorehabil Neural Repair. 2015;29(6):499–508. doi: 10.1177/1545968314554625.

40. Reisman DS, McLean H, Keller J, Danks KA, Bastian AJ. Repeated split-belt treadmill training improves poststroke step length asymmetry. Neurorehabil Neural Repair. 2013;27(5):460–468. https://pubmed.ncbi.nlm.nih.gov/23392918/. Accessed Jul 29, 2024. doi: 10.1177/1545968312474118.

41. Grasso R, Bianchi L, Lacquaniti F. Motor patterns for human gait: Backward versus forward locomotion. J Neurophysiol. 1998;80(4):1868–1885.

42. Choi J, Son S, Park S. A backward walking training program to improve balance and mobility in children with cerebral palsy. Healthcare. 2021;9(9):1191. https://www.mdpi.com/2227-9032/9/9/1191. Accessed Jul 29, 2024. doi: 10.3390/healthcare9091191.

43. Hawkins KA, Balasubramanian CK, Vistamehr A, et al. Assessment of backward walking unmasks mobility impairments in post-stroke community ambulators. Top Stroke Rehabil. 2019;26(5):382–388. https://pubmed.ncbi.nlm.nih.gov/31081491/. Accessed Jul 29, 2024. doi: 10.1080/10749357.2019.1609182.

44. Patterson SL, Rodgers MM, Macko RF, Forrester LW. Effect of treadmill exercise training on spatial and temporal gait parameters in subjects with chronic stroke: A preliminary report. J Rehabil Res Dev. 2008;45(2):221–228. https://www.ncbi.nlm.nih.gov/pmc/articles/PMC2998758/. Accessed Jul 29, 2024.

45. Hornby TG, Campbell DD, Kahn JH, Demott T, Moore JL, Roth HR. Enhanced gait-related improvements after therapist-versus robotic-assisted locomotor training in subjects with chronic stroke: A randomized controlled study. Stroke. 2008;39(6):1786–1792. https://pubmed.ncbi.nlm.nih.gov/18467648/. Accessed Jul 29, 2024. doi: 10.1161/STROKEAHA.107.504779.

46. Silver K, Macko RF, Forrester LW, Goldberg AP, Smith GV. Effects of aerobic treadmill training on gait velocity, cadence, and gait symmetry in chronic hemiparetic stroke: A preliminary report. Neurorehabil Neural Repair. 2000;14(1):65–71. doi: 10.1177/154596830001400108.

47. Middleton A, Braun CH, Lewek MD, Fritz SL. Balance impairment limits ability to increase walking speed in individuals with chronic stroke. Disabil Rehabil. 2017;39(5):497–502. doi: 10.3109/09638288.2016.1152603.

